# Clinical Features of Genetic Resilience in Chronic Obstructive Pulmonary Disease

**DOI:** 10.1101/2023.03.06.23286843

**Authors:** Auyon J. Ghosh, Matthew Moll, Jonathan Hess, Brian D. Hobbs, Angela Love, Liam Coyne, Michael H. Cho, Peter Castaldi, Frank A. Middleton, Andras Perl, Edwin K. Silverman, Craig P. Hersh, Stephen J. Glatt

## Abstract

**Introduction:** In the personalized risk quantification of chronic obstructive pulmonary disease (COPD), genome-wide association studies and polygenic risk scores (PRS) complement traditional risk factors, such as age and cigarette smoking. However, despite being at considerable levels of risk, some individuals do not develop COPD. Research on COPD resilience remains largely unexplored.

**Methods:** We applied the previously published COPD PRS to whole genome sequencing data from non-Hispanic white and African American individuals in the COPDGene study. We defined genetic resilience as individuals unaffected by COPD with a polygenic risk score above the 90^th^ percentile. We defined risk-matched case individuals as those with COPD (i.e., FEV_1_/FVC < 0.70) and a PRS above the 90^th^ percentile. We defined low risk individuals without COPD (i.e., FEV_1_/FVC > 0.70) as a polygenic risk score below the 10^th^ percentile. We compared genetically resilient individuals to risk-matched individuals with COPD and low risk individuals by demographics, lung function, respiratory symptoms, co-morbidities, and chest CT scan measurements. We also performed survival analyses, differential expression analysis, and matching for sensitivity analyses.

**Results:** We identified 211 resilient individuals without COPD, 605 genetic risk-matched individuals with COPD, and 527 low-risk individuals without COPD. Resilient individuals had higher FEV_1_ % predicted and lower percent emphysema. In contrast, resilient individuals had higher airway wall thickness compared to low-risk unaffected individuals. While there was no difference in survival between low-risk and resilient individuals, resilient individuals had higher survival compared to risk matched cases. We also identified two genes that were differentially expressed between low-risk unaffected individuals and resilient individuals.

**Conclusion:** Genetically resilient individuals had a reduced burden of COPD disease-related measures compared to risk-matched cases but had subtly increased measures compared to low-risk unaffected individuals. Further genetic studies will be needed to illuminate the underlying pathobiology of our observations.

## Introduction

Chronic obstructive pulmonary disease (COPD) is a common and highly morbid disease(1). Cigarette smoking is the primary environmental risk factor for COPD(2). However, only a minority of smokers develop COPD, with an estimated prevalence from 20% to 50% in elderly populations(3). In addition, there is marked heterogeneity in disease susceptibility and clinical manifestations of COPD among smokers having a comparable burden of cigarette smoking(4). Genetic factors are thought to account for some of the observed clinical heterogeneity in COPD(5).

The estimated heritability for COPD ranges from 37% to 50%(6, 7). Genome-wide association studies (GWAS) have found several single nucleotide polymorphisms (SNP) that are associated with COPD, though the effect sizes of each individual SNP are small(5, 8). A polygenic risk score (PRS), which summarizes the effects of SNPs across the genome, has been shown to improve prediction of COPD(9). The development of the PRS represents an important advance in the understanding of the genetic risk of COPD and opens the door to studying genetic resilience in COPD.

The concept of genetic resilience, which has been successfully demonstrated in both Mendelian diseases and complex disorders, focuses on studying unaffected individuals with high genetic risk (10, 11). The existence of individuals who smoke cigarettes and are resilient to developing COPD has been recognized for several decades(3). However, most studies on the effects of smoking in COPD focus on the risk factors associated with development and progression of disease. Fewer studies have sought to characterize individuals who demonstrate resilience to smoking. Investigators from the SPIROMICS study developed a multi-dimensional clinical definition of resilience in COPD and found that resilient individuals accounted for a modest proportion of study individuals(12). However, the prevalence and clinical features of smokers who are genetically resilient to COPD is unknown.

In the Genetic Epidemiology of COPD (COPDGene) study, we used the previously published PRS for COPD to identify smokers unaffected with COPD but at high genetic risk (resilient) and compared them to smokers with COPD. We also compared the resilient individuals to smokers at low genetic risk to further refine the characteristics specific to genetic resilience in COPD. We sought to identify clinical, physiologic, and imaging characteristics that differ between resilient individuals, genetic risk-matched individuals with COPD, and low-risk unaffected individuals. In addition, we tested for differences in whole-blood gene expression between the groups. We hypothesize that individuals who are genetically resilient to COPD will have distinct clinical and biological features compared to risk-matched individuals with COPD and low genetic-risk smokers.

## Methods

### Study Participants

COPDGene is a multicenter longitudinal observational study. COPDGene was designed to identify genetic factors associated with COPD and includes 10,198 subjects enrolled at 21 centers in the United States. Further details regarding recruitment have been previously published.(13) All participants provided written, informed consent and institutional review boards approved the study at all participating centers. We defined COPD, according to the Global Initiative for Chronic Obstructive Lung Disease (GOLD) guidelines, as post-bronchodilator forced expiratory volume in one second (FEV_1_) to forced vital capacity (FVC) ratio < 0.70. Study individuals were invited to participate in 5-year (Phase 2) and 10-year (Phase 3) follow-up visits. Study participants completed questionnaires to quantify symptom burden, a standard spirometry protocol, and chest computed tomography (CT) scans.

### Polygenic Risk Scores and Definition of Genetic Resilience

Genetic risk for COPD was quantified using the polygenic risk score developed by Moll and colleagues(9). Briefly, SNP weights were generated using the estimated effect sizes from GWASs for FEV_1_ and FEV_1_/FVC. Only variants that were genotyped or imputed with an R^2^ > 0.5 were included. The authors applied a penalized regression framework that accounted for linkage disequilibrium and created a combined polygenic risk scores as a weighted sum of the scores for FEV_1_ and FEV_1_/FVC.

We defined genetic resilience as individuals unaffected by COPD (i.e., FEV_1_/FVC > 0.70) with a polygenic risk score above the 90^th^ percentile (**Figure 1**). We defined risk-matched case individuals as those with COPD (i.e., FEV_1_/FVC < 0.70) and a PRS above the 90^th^ percentile(11). We defined low risk individuals without COPD (i.e., FEV_1_/FVC > 0.70) as a polygenic risk score below the 10^th^ percentile.

**Figure 1:**
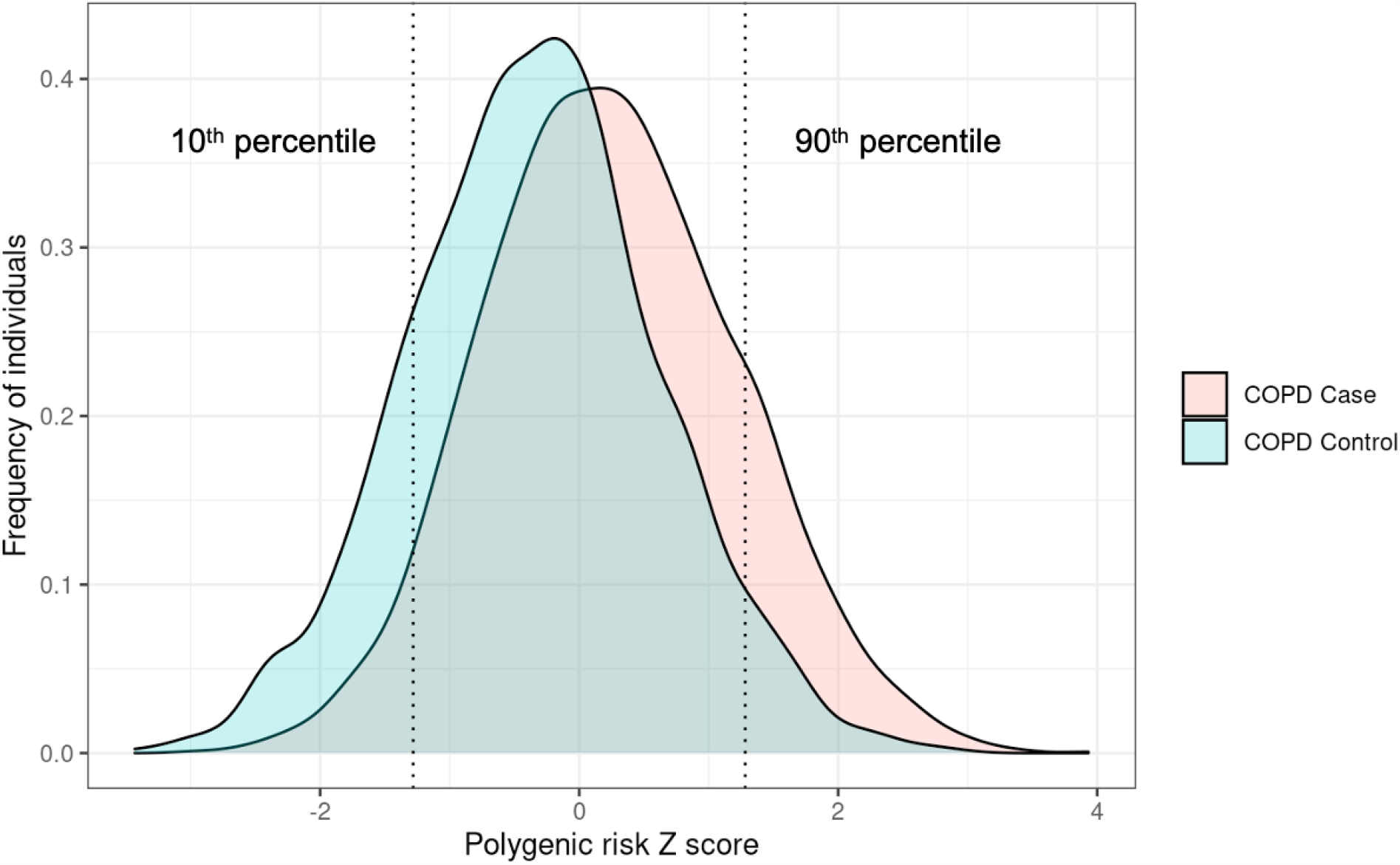
Polygenic risk score distribution in COPDGene. Polygenic risk score distribution in COPDGene, stratified by COPD diagnosis (affected individuals in salmon and unaffected individuals in light blue). Left dotted line denotes the 10^th^ percentile for polygenic risk score and right dotted line denotes the 90th percentile for polygenic risk score.

### Statistical Analysis

In race-stratified analyses, we compared genetically resilient individuals to risk-matched individuals with COPD and low risk individuals by demographics, lung function, respiratory symptoms, co-morbidities, and chest CT scan measurements, using the Student’s t test for continuous variables and Chi square test for proportions. We stratified by race given differences in principal components of ancestry. We performed multivariable regression for a subset of outcome measures, including FEV_1_ % predicted and CT scan measurements of emphysema, adjusted for age, gender, smoking pack-years, current smoking status, and the first five components of genetic ancestry. We performed survival analysis using Kaplan-Meier curves and Cox proportional hazard models, adjusted for covariates as above, using the survival and survminer packages(14, 15). Sensitivity analyses included comparison of genetically resilient individuals to risk-matched individuals with COPD after matching on age, smoking pack-years, gender, race, and current smoking status using the matchit package. Statistical analyses were performed using R (version 4.2.2).

The methods of RNA data processing have been previously published. We performed differential expression using the limma and voom packages(16, 17). Models were adjusted for age, race, sex, current smoking status, smoking pack-years, library preparation batch, and white blood cell count proportions. Surrogate variables were used to estimate other latent effects and included in the model as covariates(18). Multiple testing was controlled with a false discovery rate (FDR) < 10%.

## Results

### Comparison of genetically resilient individuals and risk-matched individuals with COPD

The COPDGene Study includes 10,198 current and former smokers. We identified 211 genetically resilient individuals (i.e., PRS ≥ 90^th^ percentile and FEV_1_/FVC > 0.70) and 605 risk-matched individuals with COPD. Compared to risk-matched cases, resilient individuals were more likely to be younger and less likely to have ever had asthma, among both non-Hispanic White (NHW) and African American (AA) individuals (**Table 1**). While NHW resilient individuals had fewer smoking pack-years but no difference in current smoking status, AA resilient individuals were more likely to be current smokers but had no difference in smoking pack years. Among both AA and NHW individuals, resilient individuals had higher FEV_1_ % predicted, less likely to have bronchodilator response, lower St. George’s Respiratory Questionnaire total score (SGRQ), and higher six-minute walk distance, compared to risk-matched individuals with COPD. Similarly, we found that resilient individuals had lower percent emphysema, defined by percent of voxels with CT attenuation less than or equal to -950 Hounsfield units on inspiratory CT, and lower Perc 15, defined as CT attenuation at the 15^th^ percentile of the lung CT histogram, across both NHW and AA individuals. In addition, NHW and AA resilient individuals had lower percent gas trapping compared to risk-matched cases. Finally, both AA and NHW resilient individuals had lower airway wall thickness, measured by Pi10, than risk-matched cases.

### Comparison of genetically resilient individuals and low-risk unaffected individuals

We identified 527 low-risk individuals (i.e., PRS ≤ 10^th^ percentile) without COPD. There was no difference in age, gender, current smoking status, smoking pack-years, or diagnosis of asthma between resilient individuals and low-risk unaffected individuals (**Table 2**). When compared to low-risk unaffected individuals, NHW resilient individuals, but not AA resilient individuals, had lower FEV_1_ % predicted. In contrast, NHW resilient individuals, but not AA resilient individuals, had higher six-minute walk distance compared to low-risk unaffected individuals. There was no difference in bronchodilator response and SGRQ between resilient and low-risk unaffected individuals in both groups. Compared to low-risk unaffected individuals, resilient individuals had lower Perc 15 but no difference in percent emphysema in both groups. NHW resilient individuals had higher percent gas trapping than low-risk unaffected individuals and there was no difference in AA resilient and low-risk unaffected individuals. Only NHW resilient individuals had higher Pi10 compared to low-risk unaffected individuals.

### Multivariable Analysis and Matching Analysis

We performed multivariable analyses for FEV_1_% predicted and percent emphysema. After adjustment for covariates, including age, gender, smoking pack-years, current smoking status, and the first five principal components of genetic ancestry, resilient individuals had higher FEV_1_ % percent predicted (NHW 41.47%, p < 0.001; AA 39.48%, p < 0.001) and lower percent emphysema (NHW -8.92%, p < 0.001; AA -4.86%, p = 0.001), compared to risk-matched cases. Compared to low-risk unaffected individuals, adjusting for the same covariates as above, NHW resilient individuals had lower FEV_1_ % predicted (NHW -6.16%, p < 0.001), while AA individuals did not. In contrast, NHW resilient individuals did not have a difference in percent emphysema while AA resilient individuals had slightly higher percent emphysema (AA 0.5%, p = 0.046) compared to low-risk unaffected individuals.

Given the imbalance in age between resilient and risk-matched case individuals, we first examined the number of individuals who were classified as resilient at initial study visit who subsequently met spirometric criteria for COPD at the 5-year follow up visit. Of the 211 individuals who were classified as resilient, 124 individuals completed the 5-year follow up visit. Among these individuals, 111 remained without COPD and only 13 met spirometric criteria for COPD. We then matched resilient subjects to risk-matched cases by nearest age and smoking pack-years and exact race, gender, and current smoking status (**Table 3**). After matching, resilient individuals still had higher six-minute walk distance, less likely to be diagnosed with asthma, lower SGRQ, higher FEV_1_ % predicted, less likely to have bronchodilator response, and a similar pattern of quantitative CT scan measures as above.

### Survival Analyses

Study individuals from COPDGene were followed for approximately 10 years. In the unadjusted model, resilient individuals had higher survival (Figure 2A). Using a Cox proportional hazard model adjusted for covariates as above, the hazard ratio (HR) for resilient individuals compared to risk-matched cases was 0.35 (95% CI 0.22 – 0.56, p < 0.001). In sensitivity analysis using the matched subset, resilient individuals had preserved higher survival (Figure 2B). In a Cox proportional hazard model adjusted only for current smoking status given the matching procedure, the HR for resilient individuals compared to risk-matched cases was 0.39 (95% CI 0.23-0.66, p < 0.001). However, in both the unadjusted model and the adjusted Cox proportional hazard model, there was no difference in survival between the resilient individuals and low-risk unaffected individuals (Figure 2C).

**Figure 2:**
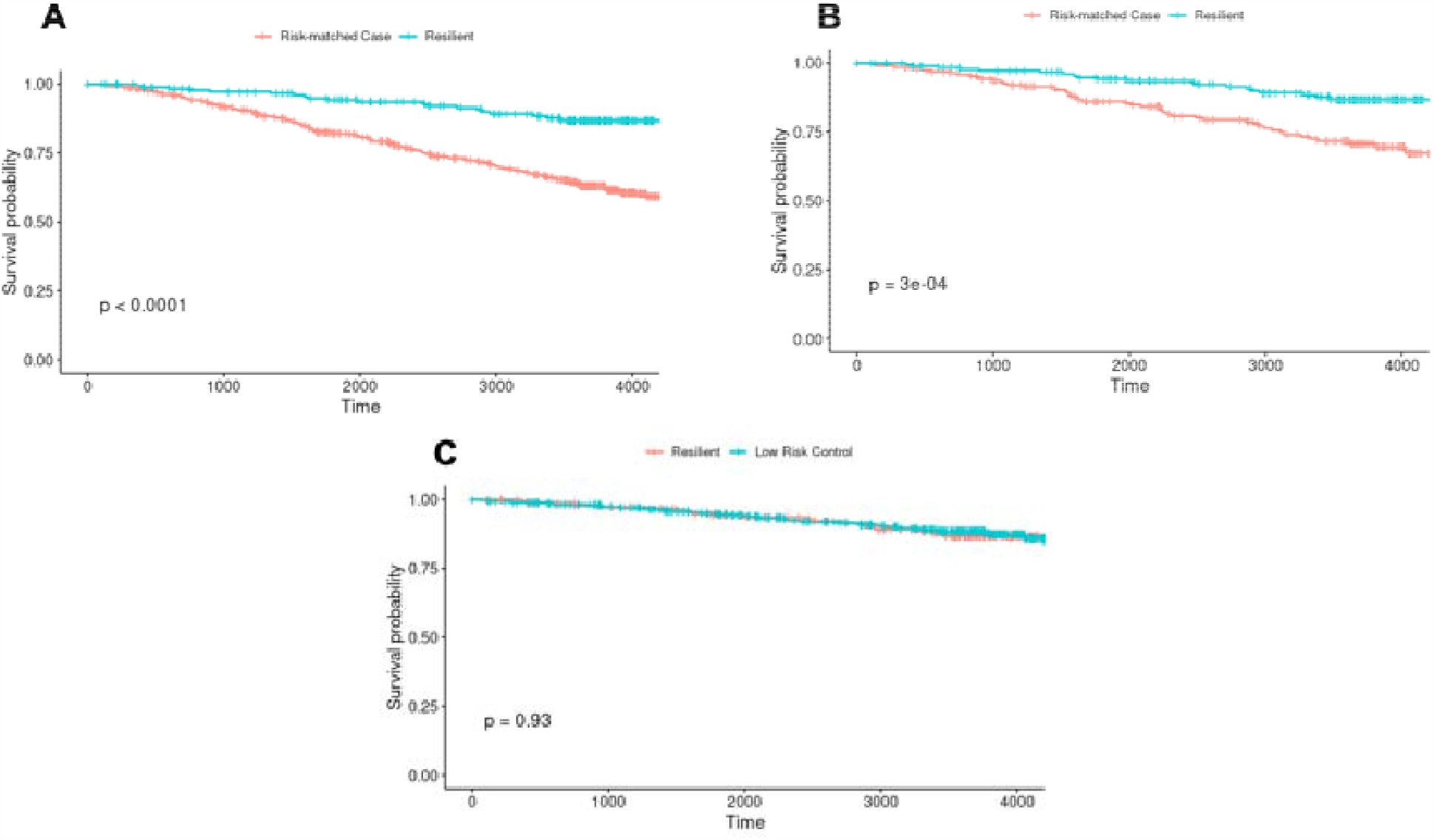
Survival in Resilient versus Risk-Match Case Individuals and Low-Risk Unaffected Individuals. Kaplan-Meier curves demonstrating the survival probability in resilient individuals (teal lines in A and B) and risk-matched case individuals (orange lines in A and B), and in resilient individuals (orange line in C) and low-risk unaffected individuals (teal line in C). (A) Survival probability in resilient individuals and risk-matched cased. (B) Survival probability in resilient individuals and risk-matched cases after matching on age, smoking pack-years, race, and gender. (C) Survival probability in resilient individuals and low-risk unaffected individuals.

### Differential Expression

There were 81 resilient individuals, 182 risk-matched cases with COPD, and 218 low-risk unaffected individuals with available whole blood RNA sequencing data from the Phase 2 visit in COPDGene. After filtering for low expression, there were 15,229 genes tested for differential expression between resilient individuals and risk-matched cases and 15204 genes tested between resilient individuals and low-risk unaffected individuals. At a 10% false discovery rate (FDR), there were no genes differentially expressed between resilient individuals and risk-matched cases. However, there were two genes, *BTN3A2* and *HLA-A*, that were differentially expressed between resilient individuals and low-risk unaffected individuals.

## Discussion

In this study, we applied a novel framework of genetic resilience to characterize individuals who are current or former smokers and have high genetic risk of COPD but appear resistant to the development of disease. We found that resilient individuals have fewer symptoms, higher functional capacity, higher lung function, and fewer radiographic signs of disease compared to risk-matched cases, even after adjusting and matching for demographic imbalances.

Interestingly, resilient individuals also were less likely to have a diagnosis of asthma. On the other hand, we found that resilient individuals have lower lung function, mixed measures of emphysema and increased airway wall thickness, but higher functional capacity compared to low-risk unaffected individuals. Finally, we have subtle but important differences in gene expression between resilient and low-risk unaffected individuals. Taken together, these findings establish genetic resilience as a distinct phenotype of individuals who smoke.

Resilience to smoking-related lung disease is of particular interest given the potential implications of identifying underlying mechanisms that could be leveraged for the development of therapeutics that could alter the trajectory of disease. By focusing on genetic resilience, as opposed to disease resilience more generally, we sought to study an important aspect of disease that is orthogonal to existing efforts to quantify resilience in COPD. While the present study represents an important advance in the application of polygenic risk and genetic resilience in COPD, we aim to further characterize the genetic variants associated with resilience in a future study.

The results of this study confirm the utility and predictive power of the polygenic risk score for COPD, with an implied positive predictive value of 74.1%. As expected, resilient individuals, by definition, did not show signs of COPD, including spirometry, functional status, and radiographic features. However, despite imbalanced demographic features, we were able to show that resilient individuals did not only appear resilient initially, but rather demonstrated resilience to disease and mortality over time. Furthermore, by contrasting resilient individuals to low-risk unaffected individuals, we were able to triangulate genetic resilience as a specific subtype beyond simply individuals unaffected by disease. Specifically, in addition to lower lung function, as expected given the nature of process underlying development of the polygenic risk score, resilient individuals also had mixed evidence of radiographic emphysema and higher airway wall thickening compared to low-risk unaffected individuals.

We have previously shown the utility of analyzing whole blood RNA sequencing to examine biological processes underlying COPD and related subtypes(19, 20). As a result of loss to follow up and studying a subset of the total study population, the differential expression analysis in this study was likely underpowered, which led to no differentially expressed genes between resilient individuals and risk-matched cases. Despite these limitations, we were able to detect two genes that were differentially expressed between resilient individuals and low-risk unaffected individuals. Both *BTN3A2* (Butyrophilin Subfamily 3 Member A2) and *HLA-A* (human leukocyte antigen alpha chain) are related to major histocompatibility complex (MHC) class I transmembrane proteins, which are essential for adaptive immune function(21, 22). This could suggest that an attenuated adaptive immune response in genetically resilient individuals may play a role in the muted reaction to cigarette smoke.

Despite many strengths, we acknowledge that there are several limitations to our study. First, our sample size was limited due to the use of a single study population and strict definition of high and low genetic risk. While we hope to expand our work to other studies in the future, we were still able to demonstrate important differences, which suggests a large true effect size of genetic resilience. Second, the polygenic risk score for COPD has indeed shown to have excellent predictive capacity for European ancestry populations, but prediction in other ancestries, including African Americans, has been less robust. Third, despite the differential expression analyses being similarly underpowered, we found modest findings that will generate hypotheses for future studies.

In summary, our study shows important clinical, radiographic, and biologic differences that separate genetically resilient individuals from similarly high-risk individuals with COPD and low-risk unaffected individuals. We demonstrate that, in addition to normal lung function, fewer symptoms, higher functional status, fewer individuals diagnosed with asthma, and decreased emphysema and airway wall thickness compared to risk-matched cases, resilient individuals also have increased airway wall thickness compared to low-risk unaffected individuals. In addition, we found that two genes associated with adaptive immunity were differentially expressed between resilient individuals and low-risk unaffected individuals. Further genetic and functional genetic studies will be required to elucidate the mechanisms that underlie these observed differences and translate these findings to novel therapies to slow and possibly prevent the development of COPD.

## Data Availability

All data produced in the present study are available upon reasonable request to the authors

## Notes

*Funding:* MM is supported by T32HL007427 and K08HL159318. BDH is supported by NIH K08HL136928 and U01 HL089856. JH is supported by NIH R21MH126494 and R01NS128535. MHC is supported by NIH R01HL137927, R01HL135142, HL147148, and HL089856. FAM is supported by NIH R01AG063499. AP is supported by NIH R01AI072648 and U01AR076092. EKS is supported by NIH R01 HL147148, U01 HL089856, R01 HL133135, R01 HL152728, P01 HL132825, and P01 HL114501. CPH is supported by NIH R01HL157879 and P01HL114501. SJG is supported by NIH R21MH126494 and R01AG064955.

COPDGene was supported by Award Number U01 HL089897 and Award Number U01 HL089856 from the National Heart, Lung, and Blood Institute. COPDGene is also supported by the COPD Foundation through contributions made to an Industry Advisory Board that has included AstraZeneca, Bayer Pharmaceuticals, Boehringer Ingelheim, Genentech, GlaxoSmithKline, Novartis, Pfizer, and Sunovion.

*Disclosures:* AJG received consulting fees from TDA Research, outside of this study. MM received grant support from Bayer. EKS received grant support from GlaxoSmithKline and Bayer. CPH reports grant support from Boehringer-Ingelheim, Novartis, Bayer and Vertex. MHC has received grant support from GlaxoSmithKline and Bayer, consulting fees from Genentech and AstraZeneca, and speaking fees from Illumina. None of the funding was allocated toward this specific work.

### Competing Interest Statement

AJG received consulting fees from TDA Research. MM received grant support from Bayer. EKS received grant support from GlaxoSmithKline and Bayer. CPH reports grant support from Boehringer-Ingelheim, Novartis, Bayer and Vertex. MHC has received grant support from GlaxoSmithKline and Bayer, consulting fees from Genentech and AstraZeneca, and speaking fees from Illumina. None of the funding was allocated toward this specific work.

### Funding Statement

This study did not receive any funding

### Author Declarations

The IRB of Brigham and Women's Hospital gave ethical approval for this work

## References

1. Global Initiative for Chronic Obstructive Lung Disease (GOLD). GOLD 2021 global strategy for the diagnosis, management, and prevention of chronic obstructive pulmonary disease.

2. Burrows B, Fletcher CM, Heard BE, Jones NL, Wootliff JS. THE EMPHYSEMATOUS AND BRONCHIAL TYPES OF CHRONIC AIRWAYS OBSTRUCTION. Lancet 1966;287:830–835.

3. Fletcher C, Peto R. The natural history of chronic airflow obstruction. BMJ 1977;1:1645–1648.

4. Castaldi PJ, Demeo DL, Hersh CP, Lomas DA, Soerheim IC, Gulsvik A, Bakke P, Rennard S, Pare P, Vestbo J, Silverman EK. Impact of non-linear smoking effects on the identification of gene-by-smoking interactions in COPD genetics studies. Thorax 2011;66:903–909.

5. Sakornsakolpat P, Prokopenko D, Lamontagne M, Reeve NF, Guyatt AL, Jackson VE, Shrine N, Qiao D, Bartz TM, Kim DK, Lee MK, Latourelle JC, Li X, Morrow JD, Obeidat M, Wyss AB, Bakke P, Barr RG, Beaty TH, Belinsky SA, Brusselle GG, Crapo JD, de Jong K, DeMeo DL, Fingerlin TE, Gharib SA, Gulsvik A, Hall IP, Hokanson JE, et al. Genetic landscape of chronic obstructive pulmonary disease identifies heterogeneous cell-type and phenotype associations. Nat Genet 2019;51:494–505.

6. Zhou JJ, Cho MH, Castaldi PJ, Hersh CP, Silverman EK, Laird NM. Heritability of Chronic Obstructive Pulmonary Disease and Related Phenotypes in Smokers. Am J Respir Crit Care Med 2013;188:941–947.

7. Palmer LJ, Knuiman MW, Divitini ML, Burton PR, James AL, Bartholomew HC, Ryan G, Musk AW. Familial aggregation and heritability of adult lung function: results from the Busselton Health Study. Eur Respir J 2001;17:696–702.

8. Hobbs BD, de Jong K, Lamontagne M, Bossé Y, Shrine N, Artigas MS, Wain L V, Hall IP, Jackson VE, Wyss AB, London SJ, North KE, Franceschini N, Strachan DP, Beaty TH, Hokanson JE, Crapo JD, Castaldi PJ, Chase RP, Bartz TM, Heckbert SR, Psaty BM, Gharib SA, Zanen P, Lammers JW, Oudkerk M, Groen HJ, Locantore N, Tal-Singer R, et al. Genetic loci associated with chronic obstructive pulmonary disease overlap with loci for lung function and pulmonary fibrosis. Nat Genet 2017;49:426–432.

9. Moll M, Sakornsakolpat P, Shrine N, Hobbs BD, DeMeo DL, John C, Guyatt AL, McGeachie MJ, Gharib SA, Obeidat M, Lahousse L, Wijnant SRA, Brusselle G, Meyers DA, Bleecker ER, Li X, Tal-Singer R, Manichaikul A, Rich SS, Won S, Kim WJ, Do AR, Washko GR, Barr RG, Psaty BM, Bartz TM, Hansel NN, Barnes K, Hokanson JE, et al. Chronic obstructive pulmonary disease and related phenotypes: polygenic risk scores in population-based and case-control cohorts. Lancet Respir Med 2020;8:696–708.

10. Chen R, Shi L, Hakenberg J, Naughton B, Sklar P, Zhang J, Zhou H, Tian L, Prakash O, Lemire M, Sleiman P, Cheng W, Chen W, Shah H, Shen Y, Fromer M, Omberg L, Deardorff MA, Zackai E, Bobe JR, Levin E, Hudson TJ, Groop L, Wang J, Hakonarson H, Wojcicki A, Diaz GA, Edelmann L, Schadt EE, et al. Analysis of 589,306 genomes identifies individuals resilient to severe Mendelian childhood diseases. Nat Biotechnol 2016;34:531–538.

11. Hess JL, Tylee DS, Mattheisen M, Børglum AD, Als TD, Grove J, Werge T, Mortensen PB, Mors O, Nordentoft M, Hougaard DM, Byberg-Grauholm J, Bækvad-Hansen M, Greenwood TA, Tsuang MT, Curtis D, Steinberg S, Sigurdsson E, Stefánsson H, Stefánsson K, Edenberg HJ, Holmans P, Faraone S V., Glatt SJ. A polygenic resilience score moderates the genetic risk for schizophrenia. Mol Psychiatry 2021;26:800–815.

12. Oh AL, Mularski RA, Barjaktarevic I, Barr RG, Bowler RP, Comellas AP, Cooper CB, Criner GJ, Han MK, Hansel NN, Hoffman EA, Kanner RE, Krishnan JA, Paine R, Parekh TM, Peters SP, Christenson SA, Woodruff PG, Anderson WH, Arjomandi M, Bhakta N, Bhatt SP, Buhr R, Curtis JL, Drummond MB, Kim V, Labaki W, Lambert A, Lazarus SC, et al. Defining Resilience to Smoking-related Lung Disease: A Modified Delphi Approach from SPIROMICS. Ann Am Thorac Soc 2021;18:1822–1831.

13. Regan E a, Hokanson JE, Murphy JR, Make B, Lynch D a, Beaty TH, Curran-Everett D, Silverman EK, Crapo JD. Genetic Epidemiology of COPD (COPDGene) Study Design. COPD J Chronic Obstr Pulm Dis 2011;7:32–43.

14. Kassambara A, Kosinski M, Biecek P, Fabian S. survminer: Drawing Survival Curves using “ggplot2.” 2019;

15. Therneau T, Lumley T. Survival Analysis. 2019;

16. Ritchie ME, Phipson B, Wu D, Hu Y, Law CW, Shi W, Smyth GK. Limma powers differential expression analyses for RNA-sequencing and microarray studies. Nucleic Acids Res 2015;43:e47.

17. Law CW, Chen Y, Shi W, Smyth GK. Voom: Precision weights unlock linear model analysis tools for RNA-seq read counts. Genome Biol 2014;15:1–17.

18. Leek JT, Johnson WE, Parker HS, Jaffe AE, Storey JD. The SVA package for removing batch effects and other unwanted variation in high-throughput experiments. Bioinformatics 2012;28:882–883.

19. Ghosh AJ, Saferali A, Lee S, Chase R, Moll M, Morrow J, Yun J, Castaldi PJ, Hersh CP. Blood RNA sequencing shows overlapping gene expression across COPD phenotype domains. Thorax 2021;doi:10.1136/thoraxjnl-2020-216401.

20. Ghosh AJ, Hobbs BD, Moll M, Saferali A, Boueiz A, Yun JH, Sciurba F, Barwick L, Limper AH, Flaherty K, Criner G, Brown KK, Wise R, Martinez FJ, Lomas D, Castaldi PJ, Carey VJ, DeMeo DL, Cho MH, Silverman EK, Hersh CP, Crapo JD, Silverman EK, Make BJ, Regan EA, Beaty TH, Castaldi PJ, Cho MH, DeMeo DL, et al. Alpha-1 Antitrypsin MZ Heterozygosity Is an Endotype of Chronic Obstructive Pulmonary Disease. Am J Respir Crit Care Med 2022;205:313–323.

21. Fagerberg L, Hallström BM, Oksvold P, Kampf C, Djureinovic D, Odeberg J, Habuka M, Tahmasebpoor S, Danielsson A, Edlund K, Asplund A, Sjöstedt E, Lundberg E, Szigyarto CA-K, Skogs M, Takanen JO, Berling H, Tegel H, Mulder J, Nilsson P, Schwenk JM, Lindskog C, Danielsson F, Mardinoglu A, Sivertsson Å, von Feilitzen K, Forsberg M, Zwahlen M, Olsson I, et al. Analysis of the Human Tissue-specific Expression by Genome-wide Integration of Transcriptomics and Antibody-based Proteomics. Mol Cell Proteomics 2014;13:397–406.

22. Crux NB, Elahi S. Human Leukocyte Antigen (HLA) and Immune Regulation: How Do Classical and Non-Classical HLA Alleles Modulate Immune Response to Human Immunodeficiency Virus and Hepatitis C Virus Infections? Front Immunol 2017;8:.

