## Supplementary material for "Clinical Features of Genetic Resilience in Chronic Obstructive Pulmonary Disease": Tables

**Table 1: Clinical Characteristics of Genetic Resilient Individuals vs Risk-Matched Individuals with COPD**

|  | Non-Hispanic White | | | African American | | |
| --- | --- | --- | --- | --- | --- | --- |
| *Characteristics* | *Risk-Matched Cases* | *Resilient Individuals* | *p value* | *Risk-Matched Cases* | *Resilient Individuals* | *p value* |
| n | 474 | 106 |  | 131 | 105 |  |
| Age, years | 63.25 (8.31) | 59.05 (8.92) | <0.001 | 57.31 (7.38) | 52.39 (5.69) | <0.001 |
| Female sex (%) | 239 (50.4) | 50 (47.2) | 0.619 | 53 (40.5) | 37 (35.2) | 0.493 |
| Current smokers | 151 (31.9) | 37 (34.9) | 0.623 | 83 (63.4) | 93 (88.6) | <0.001 |
| Smoking pack-years | 50.60 (26.93) | 34.06 (16.17) | <0.001 | 39.38 (20.47) | 35.77 (19.69) | 0.172 |
| BMI, kg/m^2^ | 28.05 (5.97) | 27.72 (4.43) | 0.588 | 28.37 (6.53) | 27.86 (5.28) | 0.515 |
| Six-minute walk distance, m | 1261.67 (407.74) | 1630.15 (297.43) | <0.001 | 1072.22 (394.27) | 1430.86 (324.02) | <0.001 |
| Asthma (%) |  |  | <0.001 |  |  | <0.001 |
| No | 283 (59.7) | 90 (84.9) |  | 74 (56.5) | 87 (82.9) |  |
| Yes | 123 (25.9) | 12 (11.3) |  | 49 (37.4) | 17 (16.2) |  |
| Don’t know | 68 (14.3) | 4 (3.8) |  | 8 (6.1) | 1 (1.0) |  |
| Age at Diagnosis | 54.57 (10.35) | 52.30 (14.70) | 0.501 | 52.52 (9.41) | 45.71 (4.57) | 0.065 |
| SGRQ Total Score | 36.69 (21.94) | 12.39 (14.59) | <0.001 | 41.21 (24.20) | 18.65 (18.99) | <0.001 |
| BODE Index | 2.74 (2.08) | 0.25 (0.63) | <0.001 | 2.90 (2.09) | 0.55 (0.90) | <0.001 |
| Percent emphysema, % | 12.73 (12.84) | 2.76 (3.22) | <0.001 | 8.02 (11.13) | 1.53 (2.60) | <0.001 |
| FEV_1_ % predicted, % | 50.51 (19.83) | 95.19 (9.18) | <0.001 | 54.76 (19.70) | 97.98 (12.40) | <0.001 |
| Bronchodilator response, % | 174 (36.7) | 9 (8.6) | <0.001 | 54 (41.5) | 9 (8.7) | <0.001 |
| Percent gas trapping, % | 39.03 (21.30) | 11.74 (10.12) | <0.001 | 29.42 (20.79) | 9.84 (9.54) | <0.001 |
| Perc15 | 62.22 (25.44) | 83.82 (17.54) | <0.001 | 79.22 (30.33) | 99.62 (23.49) | <0.001 |
| Pi10 | 2.76 (0.60) | 2.00 (0.37) | <0.001 | 2.82 (0.65) | 2.10 (0.48) | <0.001 |
| Coronary artery disease, % | 37 (7.8) | 3 (2.8) | 0.106 | 6 (4.6) | 0 (0.0) | 0.071 |
| Heart attack, % | 37 (7.8) | 1 (0.9) | 0.018 | 9 (6.9) | 2 (1.9) | 0.137 |
| Peripheral vascular disease, % | 15 (3.2) | 2 (1.9) | 0.699 | 2 (1.5) | 1 (1.0) | 1 |

Data presented as mean (SD) or no. (%). BMI: body mass index; SGRQ: St. George’s Respiratory Questionnaire; FEV_1_: forced expiratory volume in one second; Percent emphysema: percentage of lung voxels with CT attenuation ≤ -950 HU on inspiratory CT; Perc15: CT attenuation at the 15^th^ percentile of the lung CT histogram; Pi10: Square root of the wall area of a theoretical airway with internal perimeter 10mm.

**Table 2: Clinical Characteristics of Genetic Resilient Individuals vs Low-Risk Unaffected Individuals**

|  | Non-Hispanic White | | | African American | | |
| --- | --- | --- | --- | --- | --- | --- |
| *Characteristics* | *Low-Risk Unaffected Individuals* | *Resilient Individuals* | *p value* | *Low-Risk Unaffected Individuals* | *Resilient Individuals* | *p value* |
| n | 347 | 106 |  | 180 | 105 |  |
| Age, years | 59.69 (8.84) | 59.05 (8.92) | 0.517 | 53.06 (6.11) | 52.39 (5.69) | 0.362 |
| Female sex (%) | 187 (53.9) | 50 (47.2) | 0.271 | 74 (41.1) | 37 (35.2) | 0.393 |
| Current smokers | 142 (40.9) | 37 (34.9) | 0.32 | 157 (87.2) | 93 (88.6) | 0.883 |
| Smoking pack-years | 37.38 (20.05) | 34.06 (16.17) | 0.12 | 33.72 (18.28) | 35.77 (19.69) | 0.374 |
| BMI, kg/m^2^ | 28.93 (5.55) | 27.72 (4.43) | 0.04 | 28.80 (6.23) | 27.86 (5.28) | 0.196 |
| Six-minute walk distance, m | 1562.13 (301.05) | 1630.15 (297.43) | 0.042 | 1369.61 (357.66) | 1430.86 (324.02) | 0.151 |
| Asthma (%) |  |  | 0.339 |  |  | 0.141 |
| No | 310 (89.3) | 90 (84.9) |  | 145 (80.6) | 87 (82.9) |  |
| Yes | 24 (6.9) | 12 (11.3) |  | 25 (13.9) | 17 (16.2) |  |
| Don’t know | 13 (3.7) | 4 (3.8) |  | 10 (5.6) | 1 (1.0) |  |
| Age at Diagnosis | 52.90 (9.66) | 52.30 (14.70) | 0.883 | 49.90 (9.63) | 45.71 (4.57) | 0.305 |
| SGRQ Total Score | 13.76 (15.46) | 12.39 (14.59) | 0.421 | 22.31 (20.61) | 18.65 (18.99) | 0.138 |
| BODE Index | 0.26 (0.63) | 0.25 (0.63) | 0.906 | 0.63 (0.98) | 0.55 (0.90) | 0.513 |
| Percent emphysema, % | 2.24 (2.79) | 2.76 (3.22) | 0.114 | 1.07 (1.56) | 1.53 (2.60) | 0.071 |
| FEV_1_ % predicted, % | 100.85 (12.26) | 95.19 (9.18) | <0.001 | 100.10 (13.22) | 97.98 (12.40) | 0.181 |
| Bronchodilator response, % | 18 (5.2) | 9 (8.6) | 0.302 | 19 (10.6) | 9 (8.7) | 0.744 |
| Percent gas trapping, % | 9.40 (8.03) | 11.74 (10.12) | 0.026 | 7.70 (7.57) | 9.84 (9.54) | 0.07 |
| Perc15 | 90.76 (22.49) | 83.82 (17.54) | 0.005 | 108.69 (27.84) | 99.62 (23.49) | 0.007 |
| Pi10 | 1.85 (0.41) | 2.00 (0.37) | 0.001 | 2.01 (0.44) | 2.10 (0.48) | 0.122 |
| Coronary artery disease, % | 18 (5.2) | 3 (2.8) | 0.456 | 3 (1.7) | 105 (100.0) | 0.466 |
| Heart attack, % | 12 (3.5) | 1 (0.9) | 0.305 | 9 (5.0) | 0 (0.0) | 0.322 |
| Peripheral vascular disease, % | 9 (2.6) | 2 (1.9) | 0.957 | 2 (1.1) | 2 (1.9) | 1 |

Data presented as mean (SD) or no. (%). BMI: body mass index; SGRQ: St. George’s Respiratory Questionnaire; FEV_1_: forced expiratory volume in one second; Percent emphysema: percentage of lung voxels with CT attenuation ≤ -950 HU on inspiratory CT; Perc15: CT attenuation at the 15^th^ percentile of the lung CT histogram; Pi10: Square root of the wall area of a theoretical airway with internal perimeter 10mm.

**Table 3: Clinical Characteristics of Genetic Resilient Individuals vs Risk-Matched Individuals with COPD After Matching on Age, Smoking Pack-Years, Race, and Gender**

| *Characteristics* | *Risk-Matched Cases* | *Resilient Individuals* | *p value* |
| --- | --- | --- | --- |
| n | 211 | 211 |  |
| Age, years | 56.92 (7.80) | 55.74 (8.18) | 0.13 |
| Female sex (%) | 87 (41.2) | 87 (41.2) | 1 |
| Current smokers | 123 (58.3) | 130 (61.6) | 0.551 |
| Smoking pack-years | 36.42 (15.93) | 34.91 (17.99) | 0.362 |
| BMI, kg/m^2^ | 28.28 (6.49) | 27.79 (4.86) | 0.381 |
| Six-minute walk distance, m | 1221.97 (413.33) | 1530.98 (325.89) | <0.001 |
| Asthma (%) |  |  | <0.001 |
| No | 123 (58.3) | 177 (83.9) |  |
| Yes | 64 (30.3) | 29 (13.7) |  |
| Don’t know | 24 (11.4) | 5 (2.4) |  |
| Age at Diagnosis | 49.76 (8.72) | 49.59 (11.86) | 0.943 |
| SGRQ Total Score | 37.22 (24.00) | 15.51 (17.17) | <0.001 |
| BODE Index | 2.61 (2.09) | 0.40 (0.79) | <0.001 |
| Percent emphysema, % | 9.11 (12.07) | 2.16 (2.99) | <0.001 |
| FEV_1_ % predicted, % | 55.44 (20.28) | 96.58 (10.97) | <0.001 |
| Bronchodilator response, % | 84 (40.0) | 18 (8.6) | <0.001 |
| Percent gas trapping, % | 30.87 (22.08) | 10.84 (9.86) | <0.001 |
| Perc15 | 73.82 (28.19) | 91.52 (22.07) | <0.001 |
| Pi10 | 2.81 (0.66) | 2.05 (0.43) | <0.001 |
| Coronary artery disease, % | 7 (3.3) | 3 (1.4) | 0.337 |
| Heart attack, % | 15 (7.1) | 3 (1.4) | 0.008 |
| Peripheral vascular disease, % | 5 (2.4) | 3 (1.4) | 0.721 |

Data presented as mean (SD) or no. (%). BMI: body mass index; SGRQ: St. George’s Respiratory Questionnaire; FEV_1_: forced expiratory volume in one second; Percent emphysema: percentage of lung voxels with CT attenuation ≤ -950 HU on inspiratory CT; Perc15: CT attenuation at the 15^th^ percentile of the lung CT histogram; Pi10: Square root of the wall area of a theoretical airway with internal perimeter 10mm.
